# Plasma S-Adenosylmethionine is Associated with Lung Injury in COVID-19

**DOI:** 10.1101/2021.09.23.21262822

**Authors:** Evgeny Kryukov, Alexander Ivanov, Vladimir Karpov, Valery Alexandrin, Alexander Dygai, Maria Kruglova, Gennady Kostiuchenko, Sergei Kazakov, Aslan Kubatiev

**Author notes:** correspondence author: Ivanov Alexander; 125315, Russia, Moscow, Baltiyskaya str., 8.

## Abstract

**Objective:** S-Adenosylmethionine (SAM) and S-adenosylhomocysteine (SAH) are indicators of global transmethylation and may play an important role as markers of severity of COVID-19.

**Methods:** The levels of plasma SAM and SAH were determined in patients admitted with COVID-19 (n = 56, mean age = 61). Lung injury was identified by computed tomography (CT) in accordance with the CT0-4 classification.

**Results:** SAM was found to be a potential marker of lung damage risk in COVID-19 patients (SAM > 80 nM; CT3,4 vs. CT 0-2: relative ratio (RR) was 3.0; p = 0.0029). SAM/SAH > 6.0 was also found to be a marker of lung injury (CT2-4 vs. CT0,1: RR = 3.47, p = 0.0004). Interleukin-6 (IL-6) levels were associated with SAM (ρ = 0.44, p = 0.01) and SAH (ρ = 0.534, p = 0.001) levels.

**Conclusions:** High SAM levels and high methylation index are associated with the risk of lung injury in COVID-19 patients. The association of SAM and SAH with IL-6 indicates an important role of transmethylation in the development of cytokine imbalance in COVID-19 cases.

## 1. Introduction

Predictive factors and markers of severity of COVID-19 are being actively studied. Because all transmethylation reactions use S-adenosylmethionine (SAM) as a methyl group donor, SAM and its product S-adenosylhomocysteine (SAH), have a global impact on many vital processes including regulation of the expression of cytokine and inflammatory protein genes, proliferation of viral particles, among others. SARS-CoV-2 nonstructural proteins (nsp) 14 and 16, i.e., (guanine-N7)-methyltransferase (N7-Mtase) and 2’-O-methyltransferase (2’-OMTase), respectively, are SAM-binding proteins; they play a crucial role in viral transmission and viral replication [1,2]. 2’-O methylation prevents virus detection by cell innate immunity mechanisms and viral translation inhibition [3].

After giving up the methyl group, SAM is converted to SAH, and then to homocysteine (Hcy) in an SAH hydrolase (SAHH)-catalyzed reaction. The latter are biological inhibitors of transmethylation. It has also been suggested that SAM/SAH (methylation index) balance is a regulator of 2’-OMTase activity and raises the possibility that SAHH inhibitors might interfere with coronavirus replication cycle [3]. Synthetic inhibitors of N7-Mtase and 2’-OMTase are considered as promising antiviral drugs [4-6]. It was also proposed to use the restriction of the bioavailability of methionine as the main substrate for the synthesis of SAM by treating a COVID-19 patient with oral recombinant methioninase [7].

According to a recently proposed hypothesis, SARS-CoV-2 induces changes in the host’s one-carbon metabolism and methyl-group availability. Disruption of transmethylation by SARS-CoV-2 will lead to a decrease in intracellular SAM concentration. This limits the ability of cells to synthesize glutathione (GSH), a key intracellular antioxidant [8].

Notably, an increase in SAH levels and a decrease in the methylation index were considered as markers of ED both in experimental models and in individuals with chronic cardiovascular diseases [9-12]. ED is a key pathogenetic factor in lung damage in COVID-19 [13,14]. Deviation from the usual mechanisms of gene expression plays an important role in the development of ED due to changes in the activity of transmethylases which strongly depends on the levels of SAM and SAH and methylation index. The association of a high methylation index with the risk of serious lung damage suggests that ED in COVID-19 has a significant difference from that in chronic cardiovascular diseases, with respect to global methylation. Previously, an increase in plasma SAM levels was shown in a rat septic shock model [15]. To the best of our knowledge, there have been no reports on clinical studies of SAM and SAH levels in COVID-19 patients. It was previously shown that patients with sepsis had significantly higher plasma SAM and SAH than control participants and sepsis nonsurvivors had significantly higher plasma SAM and SAH levels than survivors [16]. A recent report suggested the role of high Hcy levels as a risk factor for severity or complications in COVID-19 [17,18]. Another study showed a significant correlation between Hcy levels and imaging progression on chest computed tomography (CT) from COVID-19 patients [19]. In addition, the use of a complex of B vitamins led to a decrease in the level of Hcy in COVID-19 patients; this was associated with a decrease in the period of fever and normalization of the level of D-dimer and C-reactive protein (CRP) [20].

In the present study, we aimed to analyze the plasma levels of SAM, SAH and two of their related aminothiols (Hcy and GSH) in COVID-19 patients and the possible impact of SAM and SAH on the severity of lung injury.

## 2. Methods

### 2.1. Patients

This study included 56 COVID-19 patients who were admitted in the pulmonary department of the Burdenko Main Military Clinical Hospital from September 2020 to December 2020. The study was approved by the local institutional ethics committee. Informed written consent was obtained from each patient. The reporting of this study conforms to STROBE guidelines [21].

The patients were diagnosed according to the World Health Organization’s interim guidelines for COVID-19. The main inclusion criterion was a confirmed primary coronavirus infection. Exclusion criteria included exacerbated cardiovascular disease, HIV infection, hepatitis B and C, terminal cancer, and decompensated renal failure. All patients undergoing treatment were discharged after recovery from the infection and improvement in their general condition. On admission, the patients were divided into mild, moderate, and severe condition groups according to their complaints and the results of the initial examination. On admission, patients were prescribed standard therapy in accordance with the recommendations of the Ministry of Health of the Russian Federation, which included steroids (dexamethasone, prednisolone), anticoagulants (enoxaparin, trombovazim), paracetamol (for fever > 38°C), gastroprotectant (omepaminsrazol) and vitamins B_9_+B_6_+B_12_ (angiovit), recommended 4-5h prone position and oxygen support.

Chest CT scans were performed on the 48h of patients’ admission using the Optima CT660 tomograph (GE Healthcare, USA), from the level of the thoracic entrance to the level of the diaphragm, and completed at the end of inspiration. The scanning parameters were as follows: tube voltage 120 kV, tube current 114□∼□350 mA, layer thickness 5 mm. At the end of scanning, a thin layer image with a layer thickness of 2.5 mm is automatically reconstructed and recorded as DICOM image data. The reconstruction algorithm used is with a field of view of 360 mm□×□360 mm and a matrix of 512□×□512. Image browsing and multi-plane reconstruction were performed using GE AW VolumeShare software v.4.6; images of the lungs (window width 1500, window level 500) and the mediastinum (window width 350, window level 35-40) were also observed using the same software. Image analysis was performed based on the standard protocol as described elsewhere [22]. Degree of lung damage then was assessed using the following scoring system based on percentage of lobar involvement: <5% (CT0), 5-25% (CT1), 26-49% (CT2), 50-75% (CT3) and >75% (CT4) [23]. Based on the data of an objective study of the respiratory function and blood oxygen saturation, patients were categorized into mild, moderate, and severe groups [24].

### 2.2 Laboratory procedures

On admission, venous blood was collected in 0.105 M sodium citrate tubes and centrifuged at 3000*g* for 15 min. Following that, 1.45 ml of plasma was mixed with 0.05 ml of 3 M acetic acid and the samples were frozen at -80 °C and stored until analysis.

All patients were confirmed COVID-19 positive by using SARS-CoV-2 nucleic acid detection kit “AmpliPrime® SARS-CoV-2 DUO” (Next Bio, Russia) and PCR analyzer Rotor-Gene Q (QIAGEN, Germany). Hematology analyzer MD-7600 (Meredith Diagnostics, United Kingdom), automatic biochemistry analyzers Ellipse (Analyzer Medical System, Italy), Biosen C-line (EKF Diagnostics, Germany), express immunochemiluminescent analyzer PATHFAST (Mitsubishi Chemical Medience Corporation, Japan), and erythrocyte sedimentation rate analyzer ESR 3000 (SFRI, France) were used for routine blood analysis.

GSH and Hcy levels were determined by liquid chromatography as described in a previous study [25]. SAM and SAH levels were determined by liquid chromatography – fluorescence detection as described in a previous study, with some modifications [26]. A UPLC ACQUITY system (Waters, Milford, MA, USA) was used in both these cases. Plasma (100 μl) was mixed with 5’-adenosyl-3-thiopropylamine (internal standard; 10 μl, 2.5 μM), Na-phosphate buffer (pH 8.0, 150 μl, 50 mM), and NaOH (6 μl, 1.5 M). Then, the mixture was passed through 10 mg Bond Elut PBA sorbent (Agilent, Santa Clara, CA, USA) and the phase was flushed with Na-phosphate buffer (pH 8.0, 10 mM, 0.8 ml). Analytes were eluted with HCl (0.1 ml, 0.25 M). Na-acetate (pH 5.0, 37 μl, 1 M), NaOH (9 μl, 1.5 M), and chloroacetaldehyde (20 μl, 50%) were added to the eluate for derivatization. The mixture was incubated for 4 h at 37 °C and formic acid (5 μl, 100%) was added to stop the reaction. A 5 μl aliquot of the probe sample was injected into the chromatograph.

The fluorescence signal (extinction at a wavelength of 270 nm and emission at 410 nm) was registered at a frequency of 10 s^−1^. Zorbax Eclipse plus C18 Rapid Resolution HD column (150 mm x 3 mm, 1.8 μm; Agilent, Santa Clara, USA) was used for chromatography. Flow rate was 0.37 ml/min at a temperature of 35 °C. Mobile phases were 40 mM acetic acid with 5 mM KH_2_PO_4_ + 25 μM heptafluorobutyric acid and acetonitrile. Chromatography was performed using a linear acetonitrile gradient (2%–15%) for 5 min. The column was regenerated with 50% acetonitrile for 1.5 min, and equilibrated with 2% acetonitrile for 6.5 min.

Data collection and primary processing (identification and integration of the chromatographic peaks) were performed in MassLynx v4.1 (Waters, USA). Statistical data analysis was performed using SPSS Statistics v. 22 (IBM, USA). Data on age and clinical and biochemical indicators are expressed as medians [1st; 3rd quartile]. Differences in the levels of these parameters between the patient groups were determined using rank Mann-Whitney U and Kruskal-Wallis tests. Spearman correlation coefficient (ρ) was used to describe the association between different variables. Comparison of binomial indicators (variable analysis) was carried out via relative risk (RR) and odds ratio (OR); p < 0.05 was considered to indicate a significant difference.

## 3. Results

The general characteristics of the patients are presented in Table 1. The majority (77%) of all the patients were men, and all were men in the CT3,4 group. The median patient age was 61 years, with an age range of 20–88 years. There were no smokers or regular consumers of alcohol or drugs among the patients. Most of those admitted (61%) had a mild form of COVID-19, 34% were admitted with moderate to severe infection, and 5% with severe infection. The degree of lung damage corresponded to CT4 only in two patients. Therefore, the groups CT3 and CT4 were subsequently merged. On admission, two patients underwent resuscitation/intensive therapy. A significant proportion of patients were previously diagnosed with arterial hypertension (24 out of 56, or 43%) and atherosclerosis (16 out of 56, or 29%). Significant differences were found between the patient groups in a number of laboratory indicators. In group CT3,4, there was a significant increase in erythrocyte sedimentation rate (ESR), and the levels of aspartate aminotransferase (AST), alanine aminotransferase (ALT), IL-6, CRP, SAM, and SAM/GSH. Increased levels of SAM/SAM and SAM/GSH were observed in the CT2 group.

**Table 1.** Comparative characteristics of patients with different degrees of lung damage on admission

| | CT0, 1 | CT2 | CT3,4 | $p_{\text{Kruskal-Wallis}}$ |
| --- | --- | --- | --- | --- |
| N | 26 | 16 | 14 |  |
| Age, y | 64.5 [51.3; 71.8] | 60.5 [52.8; 67.5] | 56.0 [49.3; 62.8] |  |
| Sex, man (%) | 18 (72%) | 11 (69%) | 14 (100%)<sup>†</sup> |  |
| Arterial hypertension (%) | 13 (23%) | 6 (11%) | 5 (9%) |  |
| Diabetes mellitus (%) | 3 (12%) | 2 (11%) | 2 (13%) |  |
| Atherosclerosis (%) | 9 (36%) | 3 (16%) | 4 (29%) |  |
| SpO<sub>2</sub>, % | 97 [95; 98] | 96 [94; 97.3] | 97 [96; 98] |  |
| HGB, g/l | 140 [128; 161] | 130 [119; 161] | 147 [144; 149] |  |
| HCT, % | 42 [37; 45] | 43 [36; 47] | 43 [42; 43] |  |
| RBC, 10<sup>12</sup>/l | 5.0 [4.3; 5.2] | 5.0 [3.6; 5.4] | 4.9 [4.5; 5.1] |  |
| PLT, 10<sup>9</sup>/l | 261 [216; 326] | 237 [163; 275] | 253 [210; 304] |  |
| MCV, fl | 85.5 [82.5; 88.0] | 87 [85; 90] | 86.5 [84; 91.1] |  |
| MCH, pg/cell | 30 [28.3; 30.2] | 30.5 [28.2; 31.7] | 29.6 [29.0; 30.0] |  |
| MCHC, g/l | 348 [335; 360] | 343 [329; 362] | 347 [335; 354] |  |
| WBC, 10<sup>9</sup>/l | 6.14 [4.68; 8.88] | 5.2 [3.7; 5.9] | 7.7 [4.4; 10.9] |  |
| LI | 2.4 [1.8; 3.4] | 2.9 [1.7; 4.5] | 3.6 [3.2; 5.7] |  |
| ESR, mm/h | 34 [19; 52] | 44 [18; 71] | 82 [76; 86]<sup>†‡</sup> |  |
| ALT, U/l | 28 [20; 37.5]<br>n=23 | 30.5 [21.5; 54.5]<br>n=14 | 47.5 [29.3; 142.3]<sup>‡</sup><br>n=10 |  |
| AST, U/l | 29.5 [26.8; 35.4]<br>n=24 | 29.0 [24.5; 53.3]<br>n=14 | 50.5 [37.8; 101.5]<sup>†‡</sup><br>n=10 | 0.012 |
| D-dimer, mg/l | 0.93 [0.51; 1.53]<br>n=6 | 0.71 [0.46; 0.82]<br>n=7 | 1.18 [0.72; 1.69]<br>n=7 |  |
| CRP, mg/l | 7.4 [1.8; 24.0]<br>n=25 | 32.0 [6.1; 64.8]<sup>‡</sup><br>n=15 | 71.8 [27.0; 118.5]<sup>‡</sup><br>n=11 | 0.003 |
| IL-6, ng/l | 4.85 [3.00; 18.5]<br>n=13 | 6.40 [4.39; 16.0]<br>n=9 | 14.06 [9.47; 66.40]<sup>‡</sup><br>n=11 |  |
| Ferritin, ng/ml | 226.5 [80; 324.5]<br>n=6 | 295 [211; 345]<br>n=7 | 358 [277; 764]<br>n=7 |  |
| Creatinine, $\mu\text{M}$ | 90.5 [78; 101.8] | 95.5 [86.3; 134] | 98 [85.1; 122.5] | |
| GSH, $\mu\text{M}$ | 1.81 [1.04; 2.34] | 1.15 [0.86; 1.76] | 1.22 [0.76; 1.42] | |
| Hcy, $\mu\text{M}$ | 7.4 [5.9; 9.3] | 8.3 [7.0; 10.5] | 9.1 [6.5; 12.8] | |
| SAM, nM | 59 [48; 72] | 57 [51; 84] | 84 [64; 115]<sup>‡</sup> |  |
| SAH, nM | 14.4 [4.4; 19.9] | 10.2 [8.0; 18.3] | 14.5 [12.2; 24.7] |  |
| SAM/SAH | 3.6 [2.7; 5.4] | 7.2 [4.2; 9.1]<sup>‡</sup> | 5.5 [3.3; 9.3] |  |
| SAM/GSH, nM/$\mu\text{M}$ | 32 [23; 52] | 57 [36; 131]<sup>‡</sup> | 60 [42; 285]<sup>‡</sup> | 0.017 |

<sup>‡</sup> p < 0.05 compared with “CT0,1” group
<sup>‡</sup> p < 0.05 compared with “CT2” group
ALT, alanine aminotransferase; AST, aspartate aminotransferase; CRP, C-reactive protein; CT, computed tomography; ESR, the erythrocyte sedimentation rate; GSH glutathione; HCT, hematocrit; Hcy, homocysteine; HGB, hemoglobin; IL-6, interleukin-6; LI, leukocyte index; MCH, mean erythrocyte hemoglobin; MCHC, mean corpuscular hemoglobin concentration; MCV, mean erythrocyte volume; PLT, platelets; RBC, red blood cells; SAH, S-adenosylhomocysteine; SAM, S-adenosylmethionine; SpO<sub>2</sub>, oxygenation of blood; WBC, white blood cells.

The only indicator that had a clear association with age was ferritin (ρ = 0.619, p = 0.004). In addition, its level had a significant positive correlation with ALT (ρ = 0.644, p = 0.007) and AST (ρ = 0.684, p = 0.003) levels. SAM and creatinine levels were also significantly associated with each other (ρ = 0.454, p = 0.00045). Spearman rank correlation revealed a positive association of SAM (ρ = 0.44, p = 0.01) and SAH (ρ = 0.534, p = 0.001) with IL-6 levels (Figure 1). No significant association between SAM and SAH was observed (ρ = 0.217, p = 0.108). Further, there was no significant influence of sex and age on SAM, SAH, and IL-6 levels.

**Figure 1.**
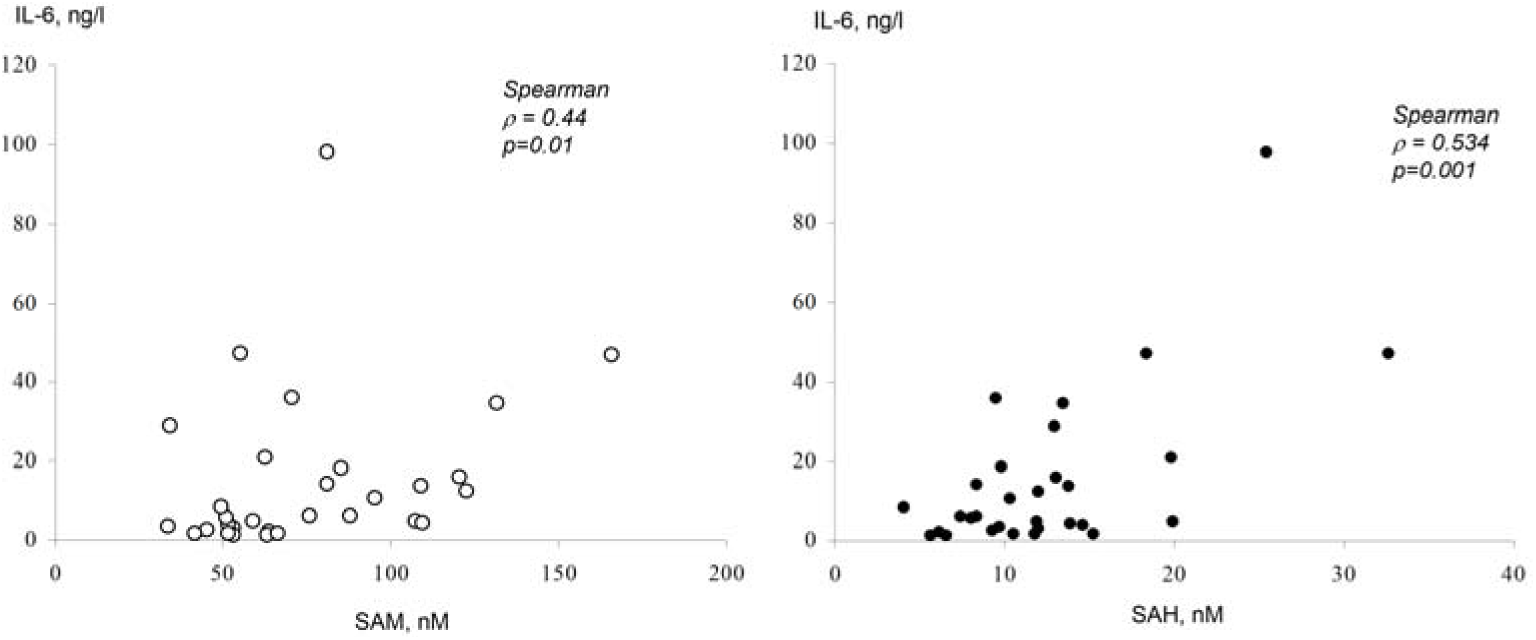
Association of SAM and SAH with IL-6 levels in COVID-19 patients

ROC analysis of our data showed that when comparing the group of patients CT0-2 with CT3,4, significant results were obtained for the following indicators: CRP (area under curve (AUC): 0.77; sensitivity – 0.727, specificity – 0.725 at cut-off 34.5 mg/l), ALT (AUC: 0.72; sensitivity – 0.6, specificity – 0.784 at cut-off 46 U/l), AST (AUC: 0.809; sensitivity – 0.8, specificity – 0.711 at cut-off 36.8 U/l) and SAM (AUC: 0.697; sensitivity – 0.714, specificity – 0.786 at cut-off 78.1 nM), see Figure 2A.

**Figure 2.**
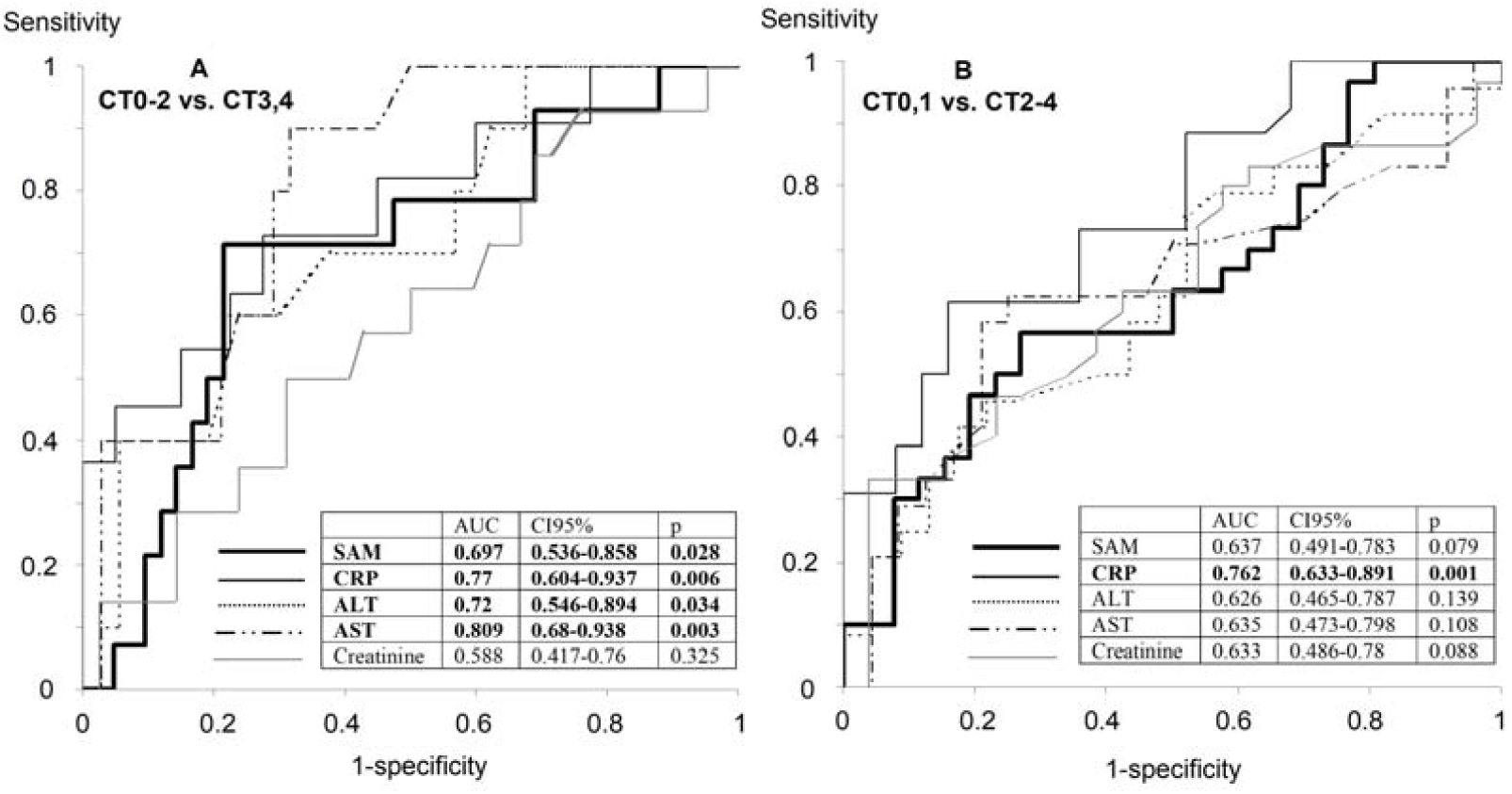
ROC analysis of the laboratory variables to compare their diagnostic performance in detecting lung injury in COVID-19. A: CT0-2 vs. CT3,4; B: CT0,1 vs. CT2-4. ALT, alanine aminotransferase; AST, aspartate aminotransferase; CRB, C-reactive protein; COVID-19, coronavirus disease 2019; CT, computed tomography; SAM, S-adenosylmethionine.

When comparing a group of patients CT0,1 with CT2-4, of all the laboratory markers used, only CRP showed relatively high results of ROC analysis (AUC: 0.762; sensitivity – 0.731, specificity – 0.64 at cut-off 13,6 mg/l), see Figure 2B. Here ALT (AUC: 0.626; sensitivity – 0.75, specificity – 0.478 at cut-off 25.5 U/l) and AST (AUC: 0.635; sensitivity – 0.542, specificity – 0.792 at cut-off 38.5 U/l) levels were not enough reliable indicators. SAM level in this case was not a sensitive enough marker (AUC: 0.637; sensitivity – 0.567, specificity – 0.731 at cut-off 66.7 nM). At the same time, the SAM/SAH ratio (AUC: 0.659, CI95%: 0.515 - 0.803, p = 0.042; sensitivity – 0.567, specificity – 0.808 at cut-off 5.88) and, especially, SAM/GSH (AUC: 0.719, CI95%: 0.584 – 0.854; p= 0.005, sensitivity – 0.767, specificity – 0.615 at cut-off 36.3 nM/μM) demonstrated relatively satisfactory performance of the ROC analysis.

The efficiency of SAM as a marker of risk of lung injury in COVID-19 patients is represented in Table 2. As shown, most of the patients (64 %) with severe lung damage (CT3,4) had SAM > 80 nM, while only 21% among the rest of the patients had SAM > 80 nM. Thus, an elevated SAM level has been associated with an increased risk of severe lung injury (CT3,4). High SAM/SAH ratio (> 6.0) and SAM/GSH (>60 nM/μM) were also found to be a marker of risk of lung damage (Table 2).

**Table 2.** Association of SAM and SAM-related indicators with the degree of lung injury of patients with coronavirus infection upon admission

| Indicator | N<sup>CT3,4</sup> | N<sup>CT0-2</sup> | RR | p | OR | 95%CI |
| --- | --- | --- | --- | --- | --- | --- |
| SAM > 80 nM | 9 of 14 | 9 of 42 | 3.0 | 0.0029 | 6.6 | 1.8-24.7 |
|  | N<sup>CT2-4</sup> | N<sup>CT0,1</sup> |  |  |  |  |
| SAM/GSH > 60 nM/μM | 15 of 30 | 5 of 26 | 2.6 | 0.017 | 4.2 | 1.25-14.1 |
| SAM/SAH > 6.0 | 20 of 30 | 5 of 26 | 3.47 | 0.0004 | 8.4 | 2.4-28.9 |

CT, computed tomography; GSH glutathione; SAH, S-adenosylhomocysteine; SAM, S-adenosylmethionine.

## 4. Discussion

Due to the close metabolic relationship, plasma SAM and SAH levels show a fairly high correlation in normal conditions [9,26,27]. However, since our study showed that COVID-19 patients with severe lung damage showed an increase in plasma SAM levels, but no significant increase in SAH levels, this clearly indicates a dysregulation of transmethylation in COVID-19.

The association of SAM levels with plasma creatinine is not surprising, since the formation of the latter requires the participation of SAM as a methyl group donor. It was previously shown that serum creatinine levels at baseline were higher in patients requiring ICU admission and mechanical ventilation and therefore this indicator found as independent risk factor for in-hospital death too [28]. Although in our work the diagnostic value of creatinine was not revealed, further study of SAM as a factor in creatinine metabolism may be of interest.

Our results are broadly consistent with previous studies of experimental endotoxinemia-induced septic shock, which showed a significant increase in plasma and liver SAM levels [15,29]. The authors suggested that this effect is due to the inhibition of transmethylases due to predominance of catabolic over anabolic processes [15]. This explains the lack of significant changes in the SAH level in this model. This is confirmed by the fact that in addition to an increase in the expression level of methionine adenosyltransferase (an enzyme that synthesizes SAM), endotoxienmia caused a significant decrease in the expression of glycine N-methyltransferase, which is the most active liver methyltransferase [29]. It is also unlikely that increase in the SAM level was due to inhibition of plasma pool utilization by the kidneys, as there was no increase in the SAH level, which is also mainly utilized through the kidneys [30]. In addition, an in vitro model of lipopolysaccharide-activated monocytes showed an increase in SAM levels on the first day, accompanied by an increase in GSH levels [31].

On the contrary, the increase in SAM levels can be considered as an adaptive reaction aimed at activating the Hcy transsulfuration pathway, leading to the synthesis of cysteine and GSH. Previously, it was demonstrated that the addition of SAM to macrophage culture attenuated the decrease in GSH levels and the expression of GSH-synthesizing enzymes, caused by the presence of lipopolysaccharide (LPS) [32]. GSH, in turn, inhibited IL-6 expression in LPS-activated alveolar macrophages [33]. However, we did not observe an increase in GSH levels in patients with high SAM levels and found no association between the levels of these metabolites. In contrast, in our study, an increase in the SAM/GSH ratio was found to be associated with an increased risk of lung damage by more than 25%, and patients with more than 50% of lung damage were characterized by both an increase in SAM and a decrease in GSH. This may indicate that the increase in the level of SAM does not play a significant protective role here.

Notably, a positive association of IL-6 with SAM and SAH was also found. IL-6 can exhibit both pro-inflammatory and anti-inflammatory properties, but an increase in its level in COVID-19 primarily plays a pro-inflammatory role, since it is an active participant in the so-called “cytokine storm” [34,35]. Numerous clinical studies show an association of elevated IL-6 levels with the severity of COVID-19, which is consistent with our results [36]. The association of IL-6 levels with the severity of lung damage in both COVID-19 and other pneumonias has been shown in previous studies [37,38].

SAM has a significant effect on the expression of IL-6, but the results of different studies are ambiguous. It was previously shown that SAM increases IL-6 production and GSH synthesis in an LPS-activated monocyte culture, but this effect is blocked by the inhibition of SAH hydrolase, an enzyme that cleaves SAH to Hcy and adenosine (Ado), or by the inhibition of methionine adenosyltransferase [39-41]. Both the above mentioned studies showed that the effect of SAM was suppressed by the inhibition of the adenosine A2 receptor. These studies concluded that the stimulation of IL-6 expression was due to an increase in the level of Ado and signaling from the A2 receptor. Ado directly caused an increase in IL-6 production in activated monocytes [40]. However, we do not yet have data on whether the increase in SAM levels is accompanied by an increase in the levels of Ado in COVID-19 patients. Indirectly, an even closer association of the levels of SAH (the precursor of Ado) with IL-6 indicates this possibility.

However, other studies on LPS-activated macrophage culture have shown that SAM significantly inhibits IL-6 expression [42,43]. This process involves the inhibition of mitogen-activated protein kinases (MAPK: ERK1/2, JNK1/2, p38) and is accompanied by an increase in global DNA methylation [42]. In addition, non-specific inhibition of DNA transmethylases suppressed this effect of SAM. Although these results may explain the association of SAM levels with IL-6 in COVID-19 patients, the association of SAH with IL-6 remains unclear, since SAH is a transmethylase inhibitor that should cause a global decrease in DNA methylation.

This study has a number of limitations. Firstly, it is a small sample size, the heterogeneity of patient groups, the lack of follow-up of patients, which makes limited generalisability of the findings in a single-center study, the findings indicate the importance of assessing SAM levels and the SAM/SAH ratio for use as markers of COVID-19 prognosis or for the use of methyltransferase inhibitors for the treatment of COVID-19.

## Conclusion

Since the methylation (capping) of viral RNA is necessary for its life cycle, the role of this metabolite in the pathophysiology of COVID-19 is not entirely clear. Elevated SAM levels can be considered as a marker of the risk of lung damage in COVID-19 patients and, most likely, a factor associated with the development of the inflammatory process. On the other hand, there are several reasons to consider an increase in SAM levels as an anti-inflammatory response of the body. The association of SAM and SAH with IL-6 suggests that they play an important role in transmethylation toward the development of cytokine imbalance in COVID-19, but more research is needed to identify the pathogenetic and therapeutic potential for correcting SAM levels.

## Data Availability

The anonymized data used to support the findings of this study are available from the corresponding author upon request.

https://figshare.com/articles/dataset/SAM_SAH_Covid_patients_data_xls/16539591

## Abbreviations

2’-OMTase: 2’-O-methyltransferase
Ado: adenosine
ALT: alanine aminotransferase
AST: aspartate aminotransferase
CRB: C-reactive protein
COVID-19: coronavirus disease 2019
CT: computed tomography
ED: endothelial dysfunction
ESR: the erythrocyte sedimentation rate
GSH: glutathione
HCT: hematocrit
Hcy: homocysteine
HGB: hemoglobin
IL-6: interleukin-6
LI: leukocyte index
LPS: lipopolysaccharide
MAPK: mitogen-activated protein kinases
MCH: mean erythrocyte hemoglobin
MCHC: mean corpuscular hemoglobin concentration
MCV: mean erythrocyte volume
N7-Mtase: (guanine-N7)-methyltransferase
Nsp: nonstructural proteins
OR: odds ratio
PLT: platelets
RBC: red blood cells
RR: relative risk
SAH: S-adenosylhomocysteine
SAHH: SAH hydrolase
SAM: S-adenosylmethionine
SARS-CoV-2: acute respiratory syndrome coronavirus-2
WBC: white blood cells

## Acknowledgments

This work was supported by the Russian Foundation for Basic Research (RFBR), project ? 20-04-60251\20.

